# Whole genome sequencing and global metabolome profiling of clinical *Mycobacterium tuberculosis* isolates provide insights to their drug resistance status

**DOI:** 10.1101/2022.09.30.22279918

**Authors:** Ashutosh Sahoo, Amit Kumar Mohapatra, Haripriya Priyadarsini, Raghuveer Varma Pemmadi, Anjan Das, Tenzin Choedon, Chaitali Nikam, Rajendra Kumar Behera, Shyam K. Masakapalli, Ranjan K. Nanda

**Affiliations:** School of Biosciences and Bioengineering, Indian Institute of Technology Mandi, Kamand, Himachal Pradesh, 175005, India; Translational Health Group, International Centre for Genetic Engineering and Biotechnology, New Delhi, 110067, India; School of Life Sciences, Sambalpur University, Sambalpur, Odisha, 768019, India; Department of Respiratory Medicine, Agartala Government Medical College, Kunjaban, Agartala, Tripura, 799006, India; Structural Biology Group, International Centre for Genetic Engineering and Biotechnology, New Delhi, 110067, India; Thyrocare Technologies, Mumbai, Maharashtra,110067, India

**Keywords:** Tuberculosis, Mycobacterium tuberculosis, drug resistance, whole genome sequencing, metabolome

## Abstract

Whole genome sequence analysis of the *Mycobacterium tuberculosis* (Mtb) isolates show correlation to their drug resistance phenotype which may also reflect in their global metabolome. In this report, clinical Mtb isolates (S1, S4, S5, S6, S7, S10) harvested from the sputum of tuberculosis patients were characterized using drug sensitive test (DST), electron microscope, whole genome sequencing (WGS) and metabolomics. Majority of these Mtb isolates showed similar size (length: 1.0–3.2 μm; width: 0.32–0.52 μm) to the H37Rv Mtb strain whereas significant variations were observed in their growth kinetics, WGS and metabolome profiles. In-silico drug resistance prediction, from the WGS data (single-nulceotide polymorphisms (SNP) pattern) of these Mtb isolates showed resistance to tuberculosis drugs and matched with DST results. Differences in the genes involved in stress response, pathogenicity, drug efflux pumps were observed between isolates but genes of the central carbon metabolic pathways and amino acid metabolism were conserved. Gas chromatography and mass spectrometry (GC-MS) based metabolite profiling of these clinical isolates identified 291 metabolites involved in various metabolic pathways and a sub set of these metabolites (glutamic acid, aspartic acid and serine) contributed to the drug resistance patterns. These clinical Mtb isolates could be useful as alternate reagent for understanding host pathogen interaction and the pipeline used for WGS analysis could be used to predict drug resistance pattern of new Mtb isolates.

## Introduction

Tuberculosis disease, caused by the infection of *Mycobacterium tuberculosis* (Mtb), affects millions of lives and is a major global issue.^1,2^ Increase in the number of multi-drug resistant tuberculosis cases in certain geographical settings is a cause of concern.^2,3^ Characterizing the clinical Mtb isolates and their drug resistance pattern, at the time of diagnosis, is critical to initiate appropriate therapeutic interventions and reduce community transmission.

However, current method of drug resistance phenotyping is carried out by culture method and it has a more than three weeks of long result waiting time.^4^ Recent advancement in whole genome sequencing (WGS) of Mtb isolates provide an alternate method for predicting drug resistance status, with certain refinement, it may reduce the result waiting period to initiate appropriate and effective personalized therapy.^5,6^

In this study, Mtb isolates from the tuberculosis patient sputum samples were characterized by their microbiological phenotyping, morphology, WGS and metabolome data. In silico analysis of the WGS data of the Mtb isolates were used to identify single nucleotide polymorphisms (SNPs) and predict drug resistance patterns. Mass spectrometry was employed to monitor the global metabolite levels of these clinical isolates and compared with reference strain like H37Rv Mtb. The adopted informatics pipeline could be useful for predicting drug resistance phenotype and the clinical Mtb isolates could be useful reagents for better understanding host pathogen interaction.

## Methodology

### Study subject recruitment and isolation of clinical Mycobacterial isolates

This study was approved by the human ethics committees of the Agartala Government Medical College, Agartala (F.4[6-9]/AGMC/Academic/IEC Committee/2015/8965, dated 25 April 2018) and International Centre for Genetic Engineering and Biotechnology, New Delhi (ICGEB/IEC/2017/07).^7^ Subjects with symptoms of tuberculosis disease like cough for 2 weeks or more, weight loss, fevere, night sweat reporting to the outpatient department of the hospital were recruited after receiving their signed informed consent forms. Sputum samples (day 0 and day 1) of these subjects were taken for sputum microscopy at hospital and culture test for drug sensitivity test using Mycobacteria Growth Indicator Tube (MGIT) in BACTEC MGIT 960 System at a National Accreditation Board for Testing and Calibration Laboratories (NABL) accredited laboratory following WHO guidelines. Clinical Mtb strains (S1, S4, S5, S6, S7, and S10) were isolated from the colonies of culture positive MGIT tubes and expand using 7H9 media supplemented with Oleic acid-albumin-dextrose-catalase (OADC, 10%), Tween-80 (0.05%, vol/vol) and glycerol (0.2%, vol/vol) at 37°C in a shaker incubator at 180 rpm in BSL-III laboratory. ^8,9^ Equal number (∼107) of each clinical isolates and laboratory Mtb strain (H37Rv) were cultured for 28 days using similar conditions and their OD was monitored at 600 nm using spectrophotometer (Biorad, USA) on alternate days for growth kinetics.^8,9^ Aliquots of these Mtb samples were pre-processed and used for further characterization using transmission electron microscope (TEM), whole genome sequencing (WGS) and mass spectrometry (MS) (**Figure 1**).

**Figure 1:**
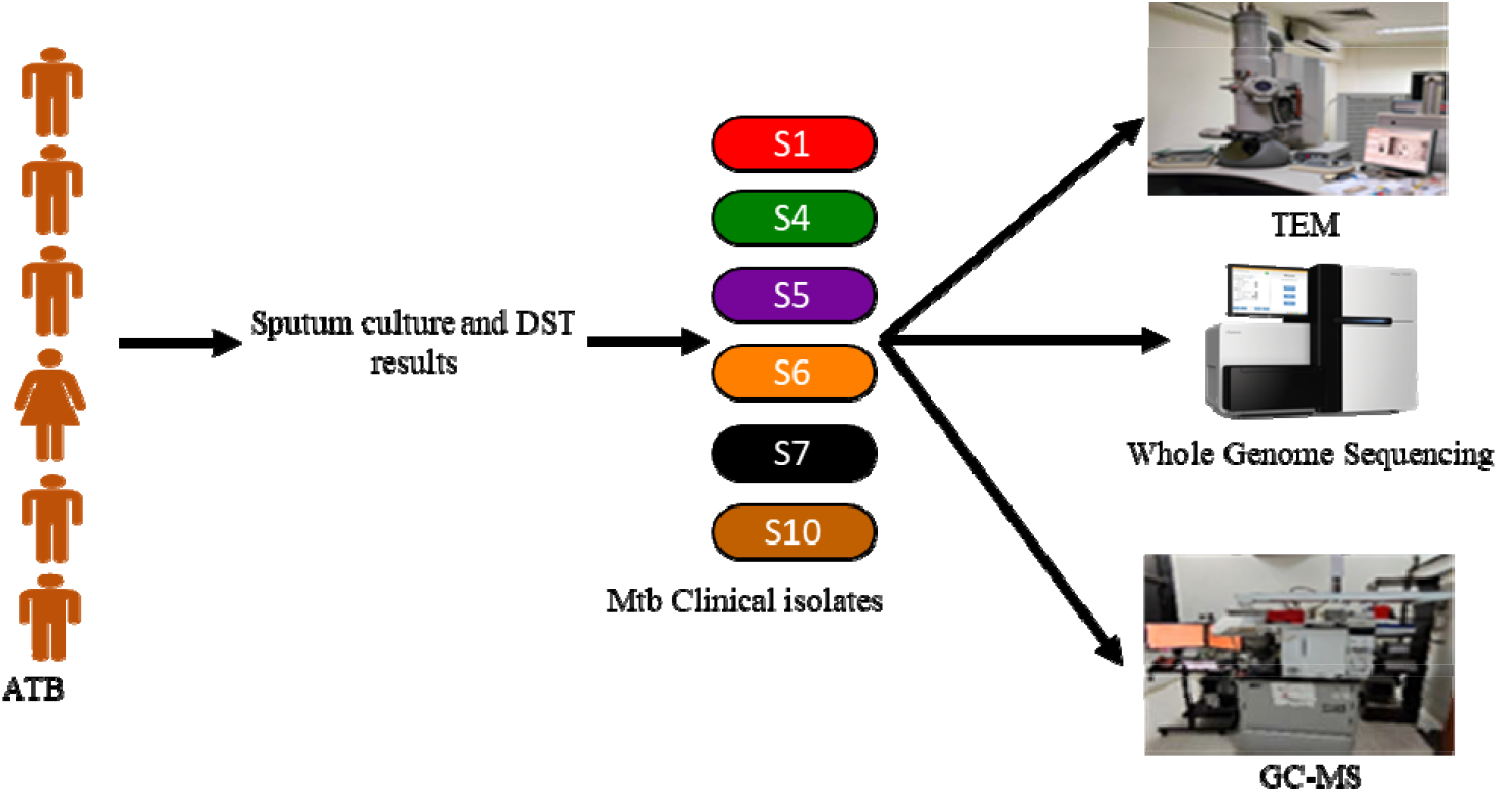
Schematics presenting the methods adopted for isolation and analysis of clinical *Mycobacterium tuberculosis* isolates. DST: drug sensitive test, TEM: transmission electron microscope, GC-MS: gas chromatography and mass spectrometry.

### Transmission electron microscopy of clinical isolates and laboratory Mycobacterial strain

Clinical isolates and laboratory Mtb strain (S1, S4, S5, S6, S7, S10, H37Rv; ∼5×10^8^ colony forming units:CFU/ml) were centrifuged at 3,000 g for 5 min. The resulting bacterial pellet was washed with phosphate buffer saline (20 mM, PBS), and incubated with Gluteraldehyde (2% in PBS:0.1 M) for 1 hour at room temperature for fixing in BSL-III laboratory.^10,11^ These bacterial preparation was washed using PBS buffer (20 mM) and twice with distilled water. After a short centrifugation, the resulting bacteria were reconstituted in distilled water (10-20 μl) before loading on a copper grid covered by carbon formvar (300 mesh, Ted Pella, USA) and stained with Uranyl Acetate (1%). The Mtb loaded grids were examined under a transmission electron microscopy (TEM, Tecnai G2 spirit, HT 120 kV) and images were captured. Using ImageJ software (vs 1.53e), the length and width of 15 individual bacteria of each samples were measured for comparative analysis.

### Global metabolite profiling of of clinical isolates and laboratory Mycobacterial strain

Five biological replicates of each Mtb samples (S1, S4, S5, S6, S7, and H37Rv), except S10 (4 replicates) were harvested at OD=1.0 at 600 nm and centrifuged at 1,500 *g* for 10 min at 4°C^12^. After washing the bacterial pellets twice with NaCl solution (0.9%) were centrifuged at 1,500 *g* for 10 min at 4°C and pre-chilled (at −80°C) Acetonitrile: Methanol: Water (40: 40: 20, 2 mL) was added.^13^ Spike in standard (Ribitol, 4 μg) was added to it and metabolites were extracted using bead beating method. Briefly, after adding Zirconium beads (0.1 mm, Biospec, CatNo. 11079101z) to the bacterial pellets, 4 cycles (on/off: 40 sec/40 sec) of bead beating was carried out. The bacterial lysates were centrifuged at 2,000 *g* for 3 min at 4°C and the supernatant was filtered (0.22 μm, nylon filters) before completely drying at 50°C using a speed vac evaporator (Labconco, USA). Toluene (100 μL) was added to each sample, again dried for at least 20 mins using speed vac at 50°C. The extracted metabolites were incubated with O-Methoxiamine HCl (40 μL, 20 mg/mL) at 60°C for 2 h at 400 rpm in a thermomixer (Eppendorf, USA). N-methyl-N-(trimethylsilyl)trifluoroacetamide (MSTFA, 70 μL) was added to the reaction mixture and incubated at 60°C for 1 h at 400 rpm in a thermomixer. Within 48 hours of derivatization, the GC-MS data acquisition using a Pegasus 4D time of flight (TOF) mass spectrometer (Leco, USA) coupled to a GC system (7890A GC from Agilent Technologies, USA) was completed.^12^ Derivatized samples (1 μL) were injected to the GC column (HP-5MS, Agilent) in splitless mode using an automatic liquid sampler (Rail system, Gestrel) and helium at a constant flow of 1 mL/min was used as the carrier gas. Separation of the metabolites were achived using a GC oven tempreture programme, from 50 to 200°C at a ramp of 8.5°C/min, from 200 to 280°C at a ramp of 6°C/min and a hold time of 5 min at final temperature. A mass range of 33 to 600 m/z at an EM voltage of 70 eV with a 20 spectra/sec scan rate and 600 sec solvent delay was selected during data acqusition. Using statistical compare feature of the ChromaTOF, all GC-MS data were analysed using parameters, minimum presence in >50% of the samples within group, signal to noise ratio (S/N)> 20, and peak width of 1.35. Analytes qualifying these criteria were aligned. Out of the identified 413 analytes, after merging peaks of similar analyte, removing silane related molecules resulted a set of 291 analytes which was selected for chemometric data analysis using Metaboanalyst 5.0.^14,15^ Data normalised using the spike in standard as the reference feature and auto-scaling was used for prinicipal component analysis (PCA) and partial least square discriminate analysis (PLS-DA) model was built. Analytes qualifying a variable important project parameter (VIP) score ≥1.8 of the PLS-DA model were selected as important. Identity of the important analytes were validated by running commercial standards using similar derivatization and GC-MS method.

### Whole genome sequencing analysis of the clinical Mycobacterial isolates and in silico prediction of drug resistance status

Genomic DNA from the clinical Mtb isolates were extracted from the exponentially growing cultures (OD=1.0 at 600 nm) following earlier reported method and quantified using Qubit 3.0 (ThermoFischer, USA)^7^. Quality of the extracted genetic material was monitored by running in agarose gel (0.8%) and TapeStation (Agilent, USA). The extracted genetic materials were sequenced using a HiSeq 4000 system (Illumina, Inc., San Diego, CA, USA) and the TruSeq SBS v3 reagent kit (2_× 150_bp) was used for library preparation. The WGS data of these clinical strains (S1, S4, S5, S6, S7, S10) are available in the NCBI database (BioProject: PRJNA595470).^7^ Additionally, sequence data of laboratory reference strains (H37Rv: NC_000962.3 and H37Ra: NC_009525.1), lineage-specific clinical isolates from India and abroad were selected from NCBI for compartive informatics analysis **(Supplementary Table 1)**. Informatics analyses included genome comparison (alignment, Average Nucleotide Identity: ANI, Single Nucleotide Polymorphisms: SNPs based phylogenetic closeness analysis and lineage classification), pathway comparison (reconstruction of metabolic pathways using RAST and BlastKOALA) and drug resistance prediction. ANI calculator was carried out employing the OrthoANIu algorithm with a minimum identity of 70%, minimum alignment length of 50 bps was used to calculate the GC contents and nucleotide identity was determined by considering the reference strain (H37Rv Mtb).^16^ Mauve-multiple Genome alignment and progressive-Mauve that uses both local alignment HODX scoring matrix to determine potential anchors from the ungapped genome sequence of selected strains was considered for complete genome alignment using H37Rv as the reference strain.^17,18^ The alignment was based on the default parameter settings that was optimised for bacterial whole genome alignment. A phylogenetic tree of the selected strains was constructed using CSI Phylogeny v1.4^19^ and visualized using FastTree tool.^20^ And SNPs qualifying with a minimum depth of SNP position to be 70×, quality >90, relative depth >10%, the minimum distance of 10 bp between SNPs and Z-score >1.96 were considered. Employing Jonny 2.1, the common SNPs in the studied clinical isolates and from the reported linage specific strains were monitored.^21^ Using Res Finder (v 4.0) the susceptibility pattern of these clinical isolates to anti-TB drugs (rifampicin, isoniazid, macrolides, fluoroquinolone, kanamycin, and clarithromycin etc.) and probable gene mutations conferring this resistance were predicted.^22,23^ A chromosomal point mutation with >60-90% and antimicrobial resistance (AMR) genes >60-90% threshold were selected and reconfirmed using Resistance gene Identifier (RGI) from CARD and compared with the microbiology drug phenotyping data.^24^

### Genome annotation and unique gene identification

The WGS Mtb sequence data were analysed employing Rapid Annotation using Subsystem Technology (RAST) with tRNAscan-SE and RNAmmer to annotate tRNA and rRNAs.^25,26^ GLIMMER2 was used for annotating coding protein sequence.^27^ The putative genes identified were verified by establishing phylogenetic context to proteins available in the FIGfam database using the BLAST against the corresponding FIGfam.^28^ RAST tool was used to classify the genes into subsystems. Orthovenn2 was used for clustering of genes among new clinical strains to segregate orthologous genes of Mycobacterium species based on similarities with reference H37Rv strain.^29^ An E-value>1e-5 and inflations >1.5 were selected for identification of homogeneous clusters and unique genes present in these clinical isolates were also identified.

### Metabolic pathway reconstruction by assigned KEGG orthology (KO numbers) to newly annotated genes

Annotated genes of all these six new clinical isolates were assigned with KEGG orthology (KO) numbers using BlastKOALA.^30^ These KO numbers were compared using G2KO with the KO numbers of the Mtb strains available in the KEGG database.^31^ Comparative pathway analysis between isolates were carried out using the KEGG mapper pathway reconstruction tool with particular focus to glycolysis, gluconeogenesis, tricarboxylic acid (TCA) cycle, pentose phosphate pathway and amino acid metabolic pathways. Using KEGG mapper, presence of the transporters including efflux pumps, conferring drug resistance and unique transporters were identified. Morpheus tool was used to identify the presence of absence of the enzymes.^32^

### Statistical Analysis

All data points were presented as mean±stdev and t-test with a P-value<0.05 was considered to be statistically significant.^33^ All statistical procedures were performed using GraphPad Prism 9.3.1.

## Results

### Characteristics of the clinical Mycobacterial strains

From the microbiology data, the clinical Mtb isolates S1, S4, S5 and S10 were found to be susceptible to rifampicin, isoniazid, ethambutol, ofloxacin, capreomycin and amikacin drugs. Whereas S6, S7 and S10 were resistant to rifampicin, isoniazid, streptomycin, clofazimine. Upon culture in 7H9 media, these clinical isolates (S1, S4, S5, S6, S7, and S10) showed slower growth kinetics compared to the virulent laboratory strains (H37Rv) **(Figure 2A)**. The clinical isolate S1 showed slower growth kinetics with a growth rate (μ) of 0.13/day, whereas the rest was in between of 0.14–0.2/day. The cells had a standard rod shape (1.0–3.2 μm in length and 0.32–0.52 μm in width), except S4, rest of the clinical Mtb isolates showed similar length and width to the laboratory H37Rv Mtb strain **(Figure 2B, 2C, 2D)**.

**Figure 2:**
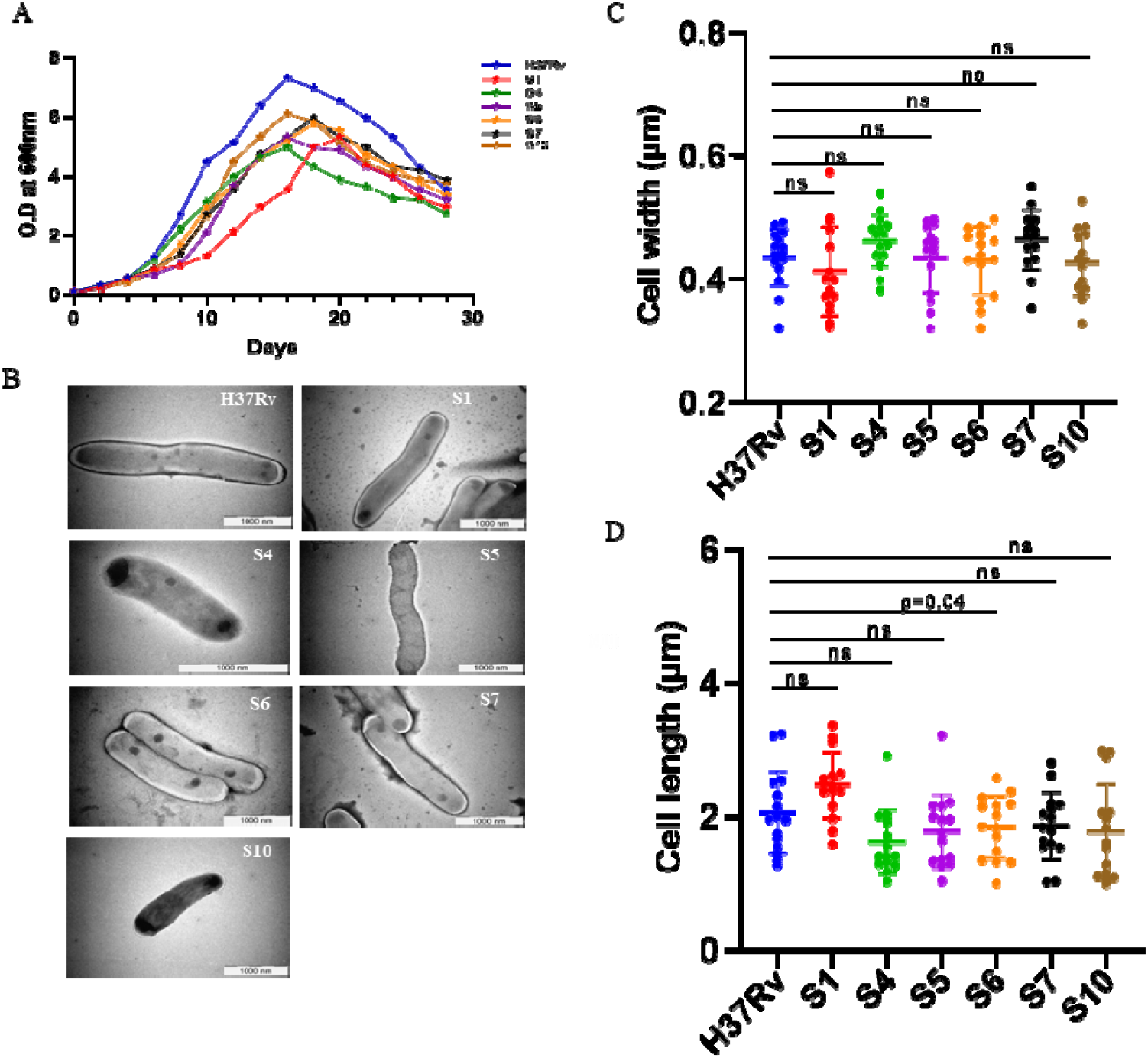
Growth kinetics and morphology of representative laboratory *Mycobactrium tuberculosis* H37Rv strain and clinical isolates showed variations. A. Growth kinetics under standard laboratory conditions. B. Transmission electron micrograph of laboratory H37Rv strain and clinical isolates (S1, S4, S5, S6, S7, S10). C. Bacterial length and D. width of individual isolates from 15 independent images. ns: not significant.

### Whole-genome sequence of the clinical Mycobacterial strains

All the raw sequences of the clinical Mtb isolates showed different genome sizes and based on fastp v0.20.0 analysis showed a coverage of 100× and pipeline employed for informatics analysis presented in **Figure 3A**.^34^ Among them S5 had a larger genome size (4,456,211 bp) followed by S10 (4,405,137 bp) and S6 (4,401,104 bp) with contig size varied from 306, 210, 124 between isolates respectively. The sequence length and contig distribution of clinical isolates (S1:4,399,078 bp:111, S4:4,396,813bp: 202, S7:4,391,229 bp: 145) showed certain difference from the other isolates **(Supplementary Table 2)**. The N50 value, ANI and SNPs of individual isolates tabulated in **Supplementary Table 2**. The GC% of all the clinical isolates was 65.6% except for S10 (65.5%). The ANI score of the S7 isolate was higher (99.79%) than other isolates compared to H37Rv reference strain. The genome sequence alignment of all clinical isolates with H37Rv showed a high degree of similarity **(Figure 3B)**. In the S5 isolate, 3148 SNPs were identified and similarly, 2494, 2519, 1950, 1280 and 2870 SNPs were observed in S1, S4, S6, S7 and S10 isolates respectively **(Supplementary Table 2)**. A set of 414 SNPs were found to be common to all study isolates and the rest were unique **(Supplementary Figure 1A)**. Using high-quality SNPs (quality score >90), a maximum-likelihood phylogenetic tree was constructed (**Figure 3C)**. The S5 and S1, S4 and S10 isolates have a distance of 0.093 and 0.0858 respectively with RGTB423 that belongs to East-African Indian lineage. S4 and S1, S10 clade shared internal nodes with a branch distance of 0.103. Larger branch length of 0.1749 between S6 and CCDC5079 isolates from the Beijing lineage was observed. The S7 isolate was grouped closer to the reported H37Rv and H37Ra reference strains with a branch length of 0.283. The SNPs distribution between isolates showed a common set of 1558 in clinical isolates (S1, S4, S5, S10) and a comparison of S6 with reported strains belonging to Beijing lineage showed 1387 SNPs **(Supplementary Figure 1B, 1C)**.

**Figure 3:**
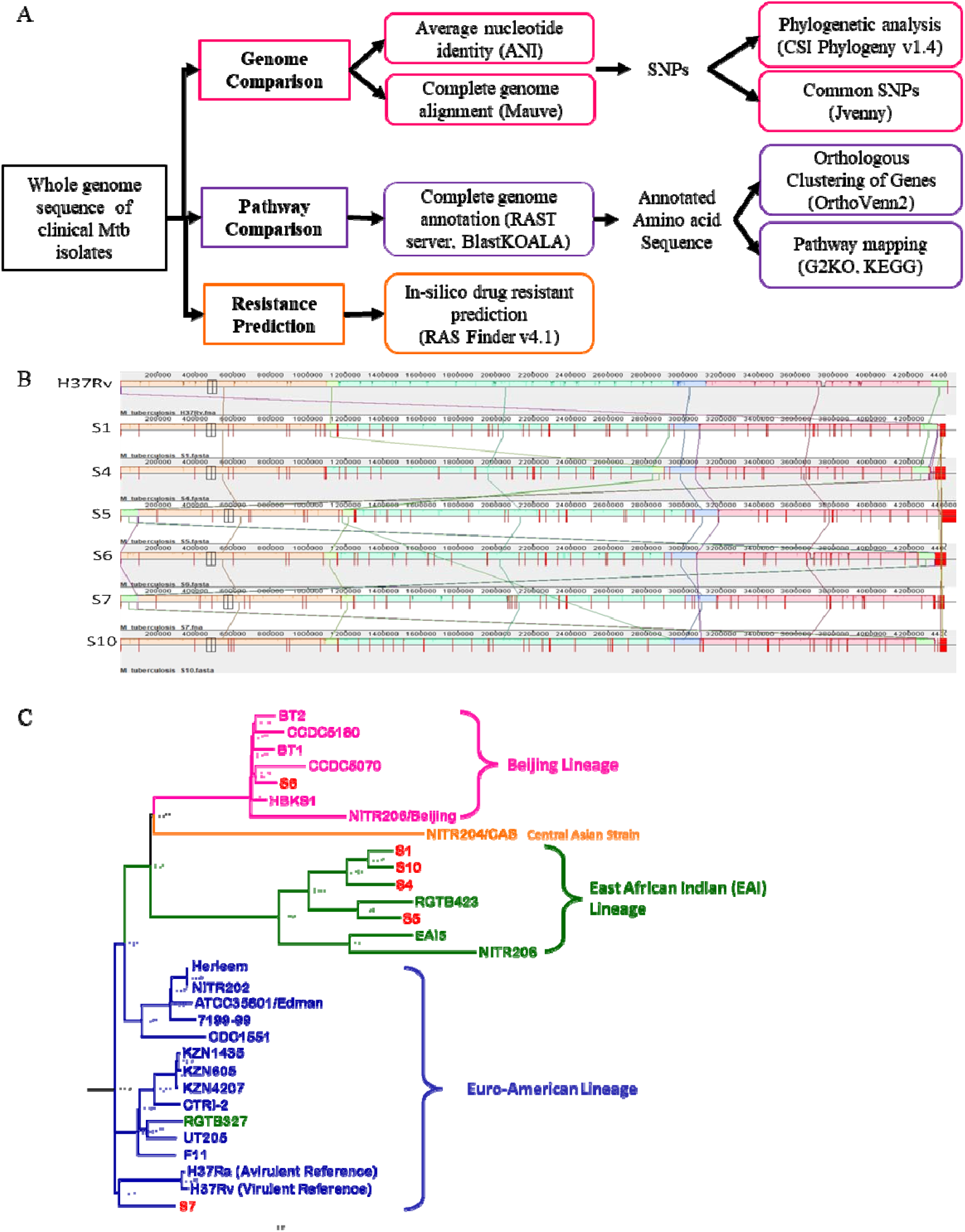
Comparative genome sequence analysis of the new clinical *Mycobacterium tuberculosis* isolates with 23 reported strains from major lineages. A. Pipeline used for comparative whole genome sequence analysis for functional characterization and drug resistance prediction. B. Complete genome alignment of new clinical isolates and H37Rv strain for nucleotide identity and single-nucleotide polymorphisms (SNPs).C. Maximum likelihood phylogenetic tree of of the clinical isolates and strains of multiple lineages constructed using CSI Phylogeny v1.4.

### Prediction of drug resistance status of the clinical Mycobacterial isolates

Reported SNPs that confer resistance to TB drugs and identified in Mtb clinical isolates presented in **Supplementary Table 3 and Supplementary figure 2**. All the mutated SNPs that become the base for predicting resistance or susceptible status in clinical isolates to known drugs like ofloxacin, capreomycin, kanamycin, amikacin, moxifloxacin, clofazimine and ethanamide were tabulated **(Supplementary Table 4)**. Based on these mutation patterns, in-silico analysis predicted resistance to rifampicin, isoniazid and streptomycin drugs in S1 and S6 isolates **(Table 1)**. The S7 isolate showed resistance to isoniazid. The predicted drug resistance status of the clinical isolates matched with the microbiological laboratory results **(Table 1)**. The total coding DNA sequences (CDS) of these clinical isolates showed variation and the annotated genes contributes to various functions **(Supplementary Table 5)**. Approximately **∼**23-24% of CDS were categorised into the virulence, disease, and defence categories with additional numbers involved in the invasion and intracellular resistance categories (**Supplementary Table 6**). Except in S6 isolate, amino acid metabolism associated genes were annotated in all the study isolates. A set of copper resistance genes and transporters were predicted to be present in S4, S5 and S6 isolates. Additional genes involved in regulation, signalling, DNA, protein, carbohydrates, fatty acids, lipids, and isoprenoids metabolism were identified in these clinical isolates (**Supplementary Table 6)**.

**Table 1:** Predicted drug resistance status of the new clinical *Mycobacterium tuberculosis* isolates from in-silico analysis and the microbiology experimental data. R/S: Resistance/Susceptibility. The matched cases were colour coded as gray and the drug resistance status is presented in black.

|  | Microbiology results |  |  |  |  |  | In-silico drug resistance prediction |  |  |  |  |  |
| --- | --- | --- | --- | --- | --- | --- | --- | --- | --- | --- | --- | --- |
| Drug Names | S1 | S4 | S5 | S6 | S7 | S10 | S1 | S4 | S5 | S6 | S7 | S10 |
| Rifampicin | S | S | S | R | S | S | R | S | S | R | S | S |
| Isoniazid | S | S | S | R | R | S | R | S | S | R | R | S |
| Ethambutol | S | S | S | S | S | S | S | S | S | R | S | S |
| Ofloxacin | S | S | S | S | S | S | S | S | S | S | S | S |
| Capreomycin | S | S | S | S | S | S | S | S | S | S | S | S |
| Streptomycin | R | S | S | R | S | S | R | S | S | R | S | S |
| Kanamycin | S | S | S | S | S | S | S | S | S | S | S | S |
| Amikacin | S | S | S | S | S | S | S | S | S | S | S | S |
| Moxifloxacin | S | S | S | S | S | S | S | S | S | S | S | S |
| Clofazimine | S | S | S | S | R | R | S | S | S | S | S | S |
| Ethionamide | S | S | S | S | S | S | S | S | S | S | R | S |
| Levofloxacin | Not Determined |  |  |  |  |  | S | S | S | S | S | S |
| Clindamycin |  |  |  |  |  |  | R | R | R | R | R | R |
| Clarithromycin |  |  |  |  |  |  | S | S | S | S | S | S |
| Erythromycin |  |  |  |  |  |  | R | R | R | R | R | R |
| Telithromycin |  |  |  |  |  |  | R | R | R | R | R | R |
| Tobramycin |  |  |  |  |  |  | R | R | R | R | R | R |
| Gentamicin |  |  |  |  |  |  | R | R | R | R | R | R |
| Dibekacin |  |  |  |  |  |  | R | R | R | R | R | R |
| Netilmicin |  |  |  |  |  |  | R | R | R | R | R | R |
| Lincomycin |  |  |  |  |  |  | R | R | R | R | R | R |

### Clustering of orthologous genes and comparative metabolic pathways in the clinical Mycobacterial isolates

A set of 3967 gene clusters consisting of 27,915 proteins were shared between clinical isolates and laboratory Mtb strains (**Figure 4A, 4B)** and isolate-specific unique gene clusters (S1:4, S4:5, S5:2 and S10:4) were observed. Metabolic pathway construction showed conserved Glycolysis, Gluconeogenesis and TCA cycles in these clinical isolates and reference Mtb strains **(Supplementary Figure 3A)**. Genes of clinical isolates involved in the Pentose phosphate pathway as well as pentose and glucuronate interconversion were found to be conserved **(Supplementary Figure 3C)**. These clinical isolates had all the enzymes required for biosynthesis and degradation of all proteogenic amino acids **(Supplementary Figure 3D)**. Certain unique enzymes or gene products were predicted to be present in these clinical isolates. Interestingly, with respect to the laboratory reference strains (H37Ra and H37Rv), many transporters were missing with few unique transporters identified in these clinical isolates which may have a role in their pathogenicity **(Figure 4C)**.

**Figure 4:**
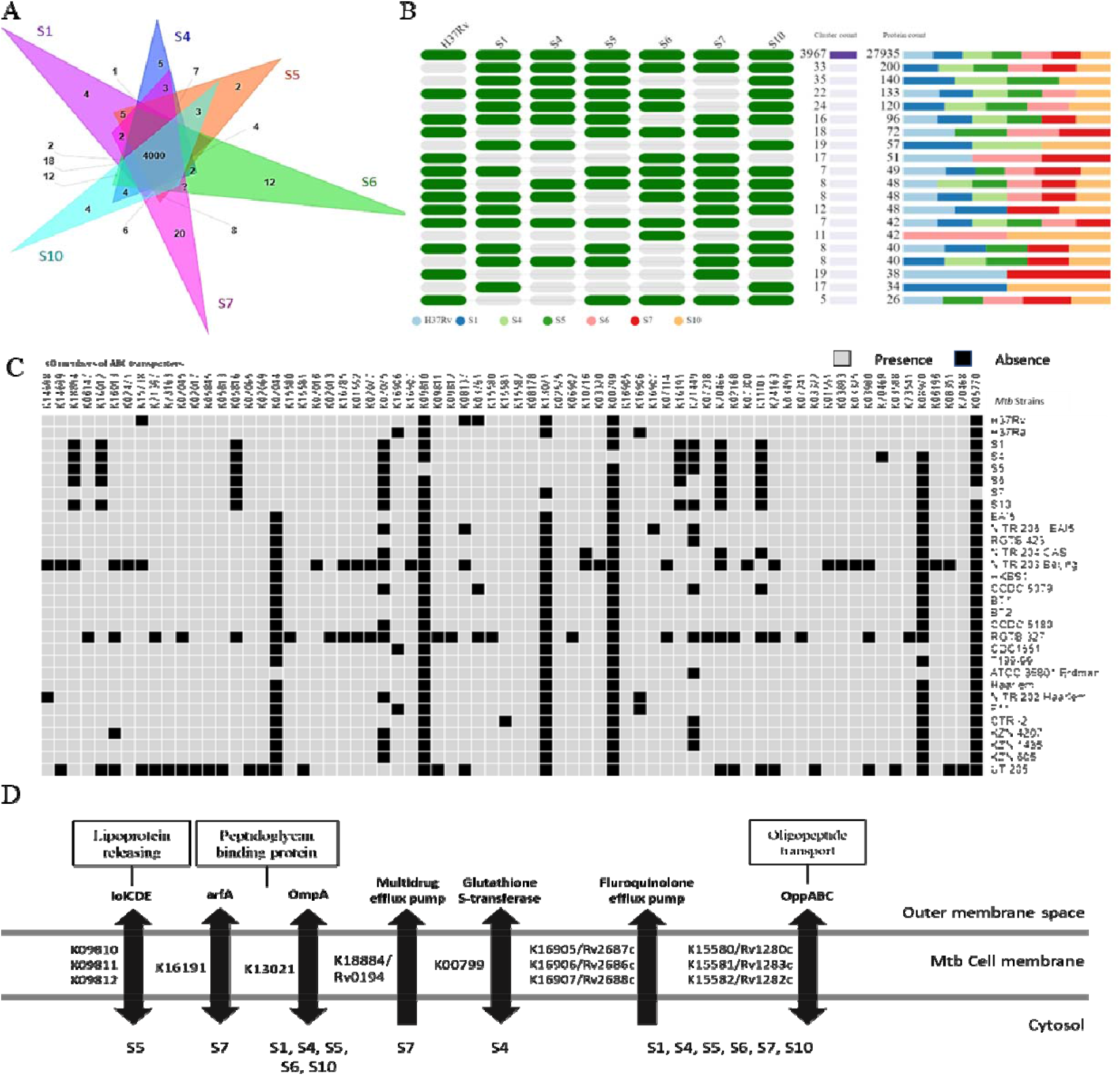
Representation of orthologous genes and unique pathways obtained from pathway comparison of the clinical *Mycobacterium tuberculosis* (Mtb) isolates. A. Venn diagram presenting orthologous clusters of annotated genes of the clinical Mtb isolates by OrthoVenn2. B. Heat map showing the proteins involved in different orthologous clusters between clinical isolates and reported Mtb strains. C. Presence and absence of the ABC transporters in clinical Mtb isolates and reported strains. D. Presence of important transporters in the newly isolated clinical Mtb isolates.

### Global metabolite profile of these clinical ioslates showed significant differences

Metabolome of the mycobacterial isolates and laboratory strain were analyzed using GC-MS after derivatization **(Figure 5A)**. Important to mention that the global metabolite analysis of the laboratory virulent Mtb H37Rv strain clustered it away from all the clinical isolates. PCA plot showed that metabolites of S1 isolate cluster separately from the other clinical isolates and laboratory reference H37Rv strain **(Supplementary Figure 4)**. PLS-DA plot showed overlap between S6 and S7 as well as S4 and S5 **(Figure 5B)**. S4 and S5 of East-African Indian lineage showed lower abundance of aspartic acid and glutamic acid with respect to S1 and S10. S4 showed a low glutamic acid level with lower aspartic acid level in S5. In the S6 isolate of Beijing lineage, serine abundance was the highest among all the clinical isolates. S7 of Euro American lineages showed lower glutamic acid and aspartic acid levels with respect to H37Rv. Glutamic acid level was found to be lower in S4 with highest in H37Rv strain. Aspratic acid level was higher in S1 isolate with a minimum level in S7. Serine levels in S6 clinical isolate was higher with very low level in S10 **(Figure 5C)**.

**Figure 5:**
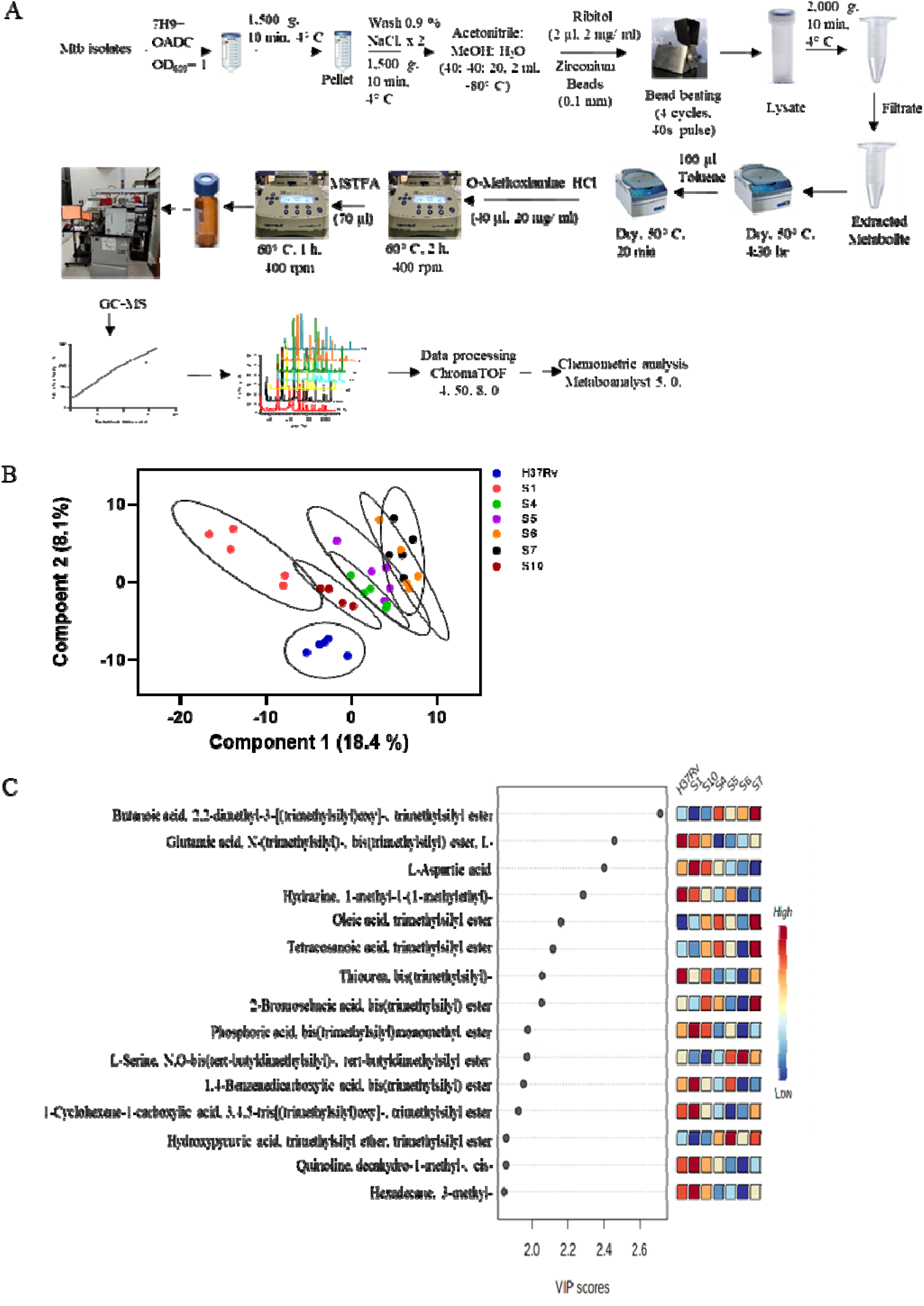
Global metabolite profiling of laboratory *Mycobacterial tuberculosis* strain (H37Rv) and clinical isolates (S1, S4, S5, S6, S7, S10) using Gas chromatography-Mass spectrometry showed significant differences. A. Pipeline used for metabolite extraction and analysis of *Mycobacterial* samples. B. Partial least squares discriminant analysis (PLS-DA) score plot showing differences in the metabolite patterns between clinical isolates and H37Rv strain. C. Variable importance in projection (VIP) score plot generated using MetaboAnalyst 5.0 showing the important deregulated metabolites.

## Discussion

Post COVID19, tuberculosis disease related cases are increasing including the number of drug resistant cases. Appropriate personalized treatment to these patients is the need of the hour and providing the drug resistance pattern in shorter time span will introduce personalized therapeutic interventions to cure the patients. Mtb the pathogen causing tuberculosis is divided into multiple lineages and show varied drug resistance patterns, virulence and transmission dynamics.^35^ Variation in virulence of these Mtb isolates from clinical materials showed dependence to patient immunity, degree of exposure and other factors including environment. It is critical to characterize clinical Mtb isolates at multiple molecular levels like WGS and their metabolic components to rationally predict their drug resistant patterns.

The difference in the growth kinetics observed in these clinical isolates could be partially explained to the lack of adoption to the available carbon and nitrogen components present in the 7H9 media. These clinical isolates, irrespective of their drug resistance and susceptible status, showed similar size with the laboratory H37Rv strain. One of the clinical isolate S4 was found to be smaller in size then the rest of the Mtb isolates. Similar morphological parameters were reported from clinical Mtb isolates from African, South east Asian and Russina tuberculosis patients. As expected the average whole genome size of the Mtb isolates was ∼4.4 Mbps with a high GC content (65.5-65.6%). The genome sequence of these clinical strains were compared with available drug susceptible and resistant Mtb strains reported from East Asia, Africa, Russia, North America and East Europe. A set of 414 SNPs were found to be common among the Mtb clinical isolates. These SNPs distribution provided insights into the evolutionary pattern of these new clinical isolates. In the study clinical site, circulating Mtb isolates of East African-Indian (S1, S4, S5, S10), Beijing (S6) and Euro-American lineages (S7) were observed. Hence identifying the genomic markers, in clinical isolates, which could predict the drug resistance pattern might rationalise the therapeutic modalities.^36^

SNP clustering analysis of all clinical Mtb isolates (S1, S4, S5 and S10), of Beijing lineage (S6, CCDC5079, CCDC5180, BT1, BT2) and clinical isolates of EAI lineage strains showed similarity of ∼30% (1387) and ∼34% (1558) SNPs respectively. These clustered SNPs among lineages of Mtb strains shows their adaptation, evolutionary changes and mutation conferring resistance to tuberculosis drugs. In silico prediction of the drug resistance patterns of the S1 and S6 clinical isolates showed resistance to rifampicin, isoniazid, and streptomycin and susceptibile to capromycine and mofloxcine corroborating culture test results. However, SNP profile of S6 isolate showed resistance to ethambutol and did not match to the microbiology data. These observations indicate that the algorithms for predicting drug resistance patterns of clinical Mtb isolates need further refinement and provide useful clinically revelant information.^37^

In the clinical drug resistance isolates few unique proteins were identified which may be partially involved in conferring drug resistance status. For example, EsxL (Rv1198) protein down regulating MHC-II by suppressing the nitric oxide production and p38 MAPK pathway in the macrophages and helps drug resistance Mtb to remain undetected for longer time for a delayed immune response.^38^ Genome annotation in these clinical isolates, predicted some genes with increased copy numbers, belonging to fatty acid synthesis and utilization, DNA metabolism and stress responses. Such alterations reported to help drug resistant Mtb to survive in the limited nutients and extreme stress condition for long.^39,40^ Interestingly, the proteins involved in the central carbon metabolism (glycolysis, gluconeogenesis, TCA cycle and Pentose phosphate pathways) were conserved in these clinical isolates irrespective of their drug resistance status. Also the genes involved in amino acid metabolism in these clinical isoaltes were found to be conserved. This suggests that possibly all these pathogens survives in the host system by maximum utilization of different carbon sources like glucose and fatty acids.^41^ Previous studies showed that fatty acids taken from the host gets utilized via TCA cycle for further gluconeogenesis to fulfil energy demand and production of glucogenic amino acids.^42^

The growth and replication of the pathogen depend on the nutritional niche of the host cell and the transporters across the membrane play a significant role. ABC transporters act as efflux pumps expelling antimicrobial agents to confer drug resistance to the pathogen.^43,44^ Comparative genomic analysis revealed presence of ABC transporters (Rv0194, Rv2686c, Rv2687c, Rv2688c) and unique ones (OmpA, Glutathion-s-transferase, opp transportors) in these clinical isolates. Rv0194, is an ATP binding cassette transporter provide resistance to ampicillin, streptomycin, and chloramphenicol and the others (Rv2686c, Rv2687c, Rv2688c) provide resistance to fluoroquinolones **(Figure 4D)**.^45,46^ Similarly, peptidoglycan binding protein transporters (ArfA and OmpA) which act as porins to maintain pH by secreting ammonia was identified in these clinical isoaltes.^47^ OmpA gene was annotated in S1, S4, S5, S6 and S10 whereas arfA was present in the S7 strain. The oppA, oppB and oppC transporters, import oligopeptides in the cell and function as oligopeptide permease and were identified in all these clinical isolates **(Figure 4D)**.^48,49^ Interestingly, S4 isolate has glutathione-S-transferase, a known detoxification enzyme involved in biodegradation of xenobiotics, protection against chemical agents and possibly provide antimicrobial.^40,50^

These clinical isolates, with varied drug resistance status and linegaes (Beijing, East African Indian and Euro American) may reflect in their metabolite phenotypes. S1 strain exhibited distinct metabolic phenotype from the rest. The clinical isolates clustered together based on their drug sensitive (S4 and S5) and resistance status (S6 and S7) in the PLS-DA plot. S4 and S5 belongs to the East-African Indian lineage showed lower aspartic acid and glutamic acid respectively than its closely linked isolates (S1 and S10). We observed slow growth rate in S1 isolate and their intracellular serine level was low which may partially explain isolate specific observed phenotype. S6, belongs to the Beijing lineage and showed high serine levels whereas S7 of Euro American lineages showed lower glutamic acid and aspartic acid levels with respect to H37Rv. Serine biosynthesis follows a multistep reaction using 3-phosphoglyceric acid (PGA) as a precursor produced during glycolysis and is essential for the survival of Mtb. Phosphoserine trasaminase serC (Rv0884c) catalyses the addition of nitrogen to the backbone of pyruvate.^51^ Reduced serine production contributes to the slow growth phenotype of Mtb and serine supplementation accelerates the growth of Mtb.^12,51,52^ Earlier reports showed lower aspartic acid level and a higher serine levels in the hypervirulent Mtb strain of Beijing lineage than a hypovirulent strain.^53^ This may indicate that S7 might show hyper-virulence than H37Rv which needs experimental validation. Glutamic acid content of H37Rv was reported to be much higher than nine Mtb strains and we also observed similar findings.^54^

In this study, we characterized clinical Mtb isolates and employed whole genome sequencing and in silico analysis to annotate presence of SNPs to predict and compare drug resistance status with microbiology findings. Interestingly, these Mtb isolates, irrespective of their drug resistance or lineage status showed varied metabolic phenotypes. This adopted informatics pipeline and metabolite data could be useful as a resource to predict drug resistance phenotype of the new clinical isolates.

## Data Availability

The raw reads of the whole-genome sequence of the clinical M. tuberculosis isolates (S1, S4, S5, S6, S7, S10) are available in the NCBI database (BioProject: PRJNA595470).

## Acknowledgment

This work was supported by the Department of Biotechnology, Government of India (grant BT/PR23238/NER/95/636/2017 to A. Das, and R. K. Nanda).

## Notes

### Competing Interest Statement

The authors have declared no competing interest.

### Funding Statement

This work was funded by the Department of Biotechnology, Government of India (grant BT/PR23238/NER/95/636/2017 to A. Das, and R. K. Nanda).

### Author Declarations

Ethics committee of Agartala Government Medical collage and International center for Genetic Engineering and Biotechnology, NewDelhi has approved this study

