## Supplemental file 29 09 2022 for "Whole genome sequencing and global metabolome profiling of clinical *Mycobacterium tuberculosis* isolates provide insights to their drug resistance status"

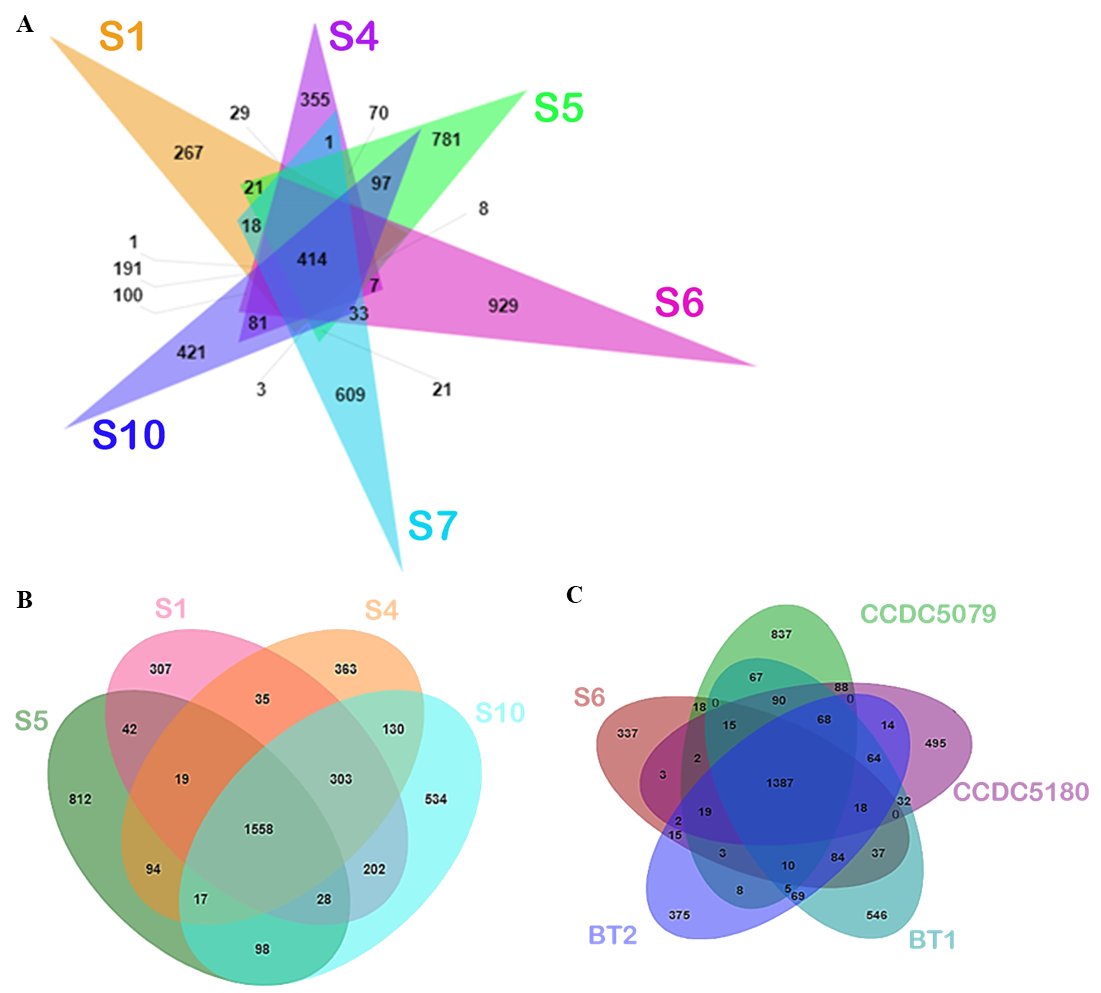

***Supplementary Figure 1:*** Distribution of single nucleotide polymorphism (SNPs) patterns in the newly identified *Mycobacterial tuberculosis* clinical isolates and reported strains from respective lineages. A. Clustering of SNPs among the *Mycobacterial tuberculosis* clinical isolates based on observed phylogenetic closeness in maximum likelihood tree using the tool Jvenn (Bardou et.al.,2014). B. Common and strain-specific SNPs among EAI linage clinical strains. C. Venn diagram showing common SNPs among Beijing lineage strains with new clinical S6 isolate.

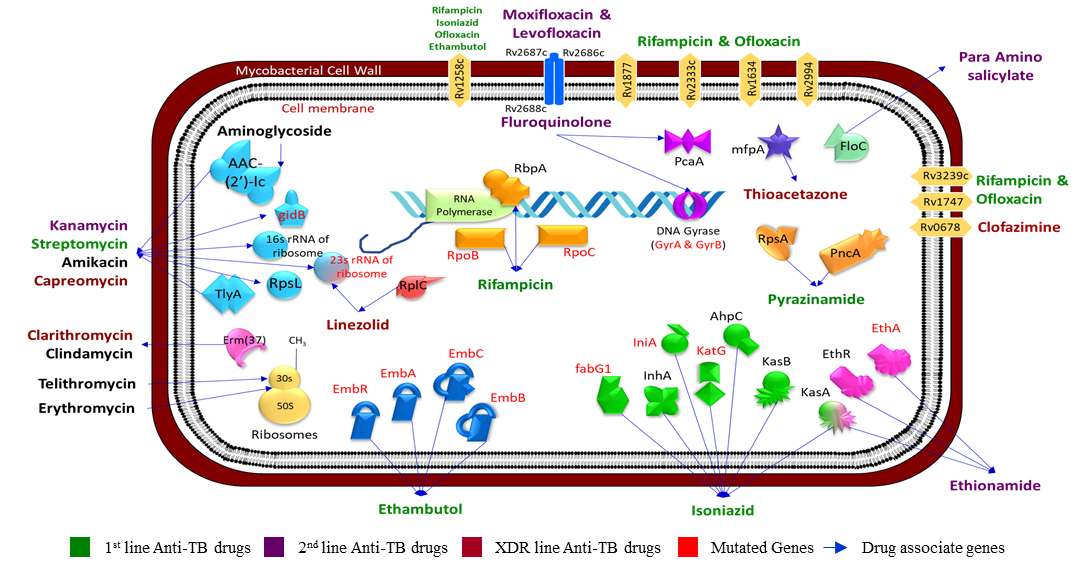

***Supplementary Figure 2:*** Model of *Mycobacterium tuberculosis* showing drug resistance genes, different classes of anti-tuberculosis and their target of action. The detailed function and drug resistance mechanism of each protein were described in supplementary Table 2.

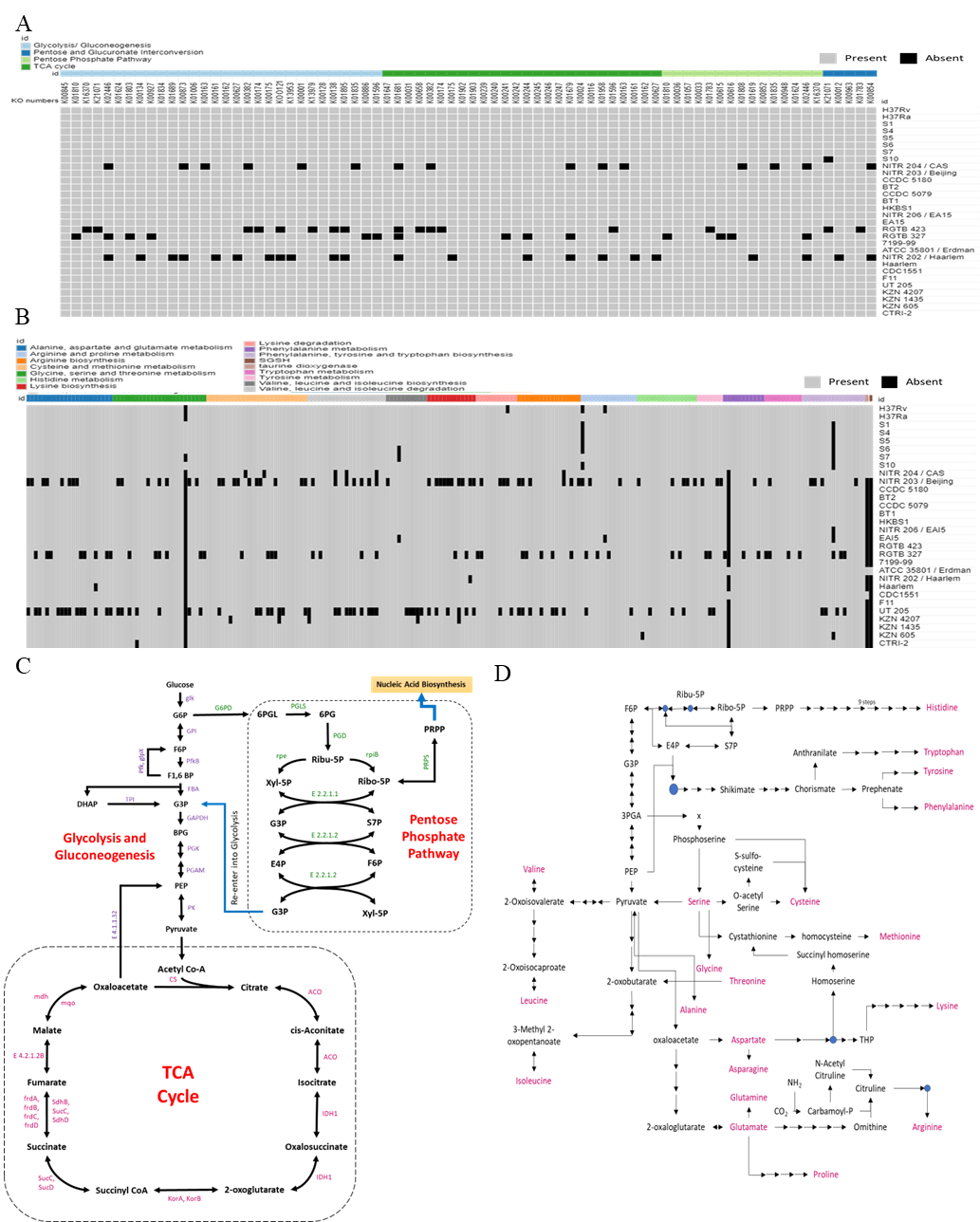

***Supplementary Figure 3:*** Pathway map of Central Carbon Metabolism and Amino acid metabolism of the identified *Mycobacterial tuberculosis* clinical isolates. A. Heatmap showing the presence or absence of the enzymes involved in Central carbon metabolism in these clinical isolates. B. Metabolic Enzymes showed their involvement in degradation and biosynthesis of proteogenic amino acids in these clinical isolates. C. Genes involved in the Glycolysis, Gluconeogenesis, TCA cycle and Pentose phosphate pathway were conserved in all these clinical isolates. D. Conserved enzymes involved in amino acid degradation as well as biosynthesis from intermediates of central carbon metabolism in *Mycobacterial tuberculosis* clinical isolates.

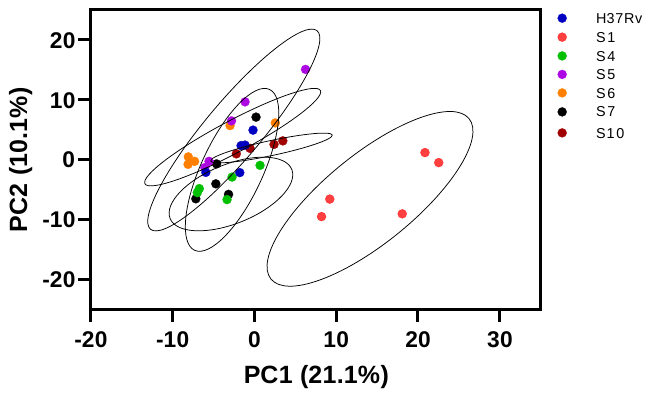

***Supplementary Figure 4*:** Score plot obtained from the Principal Component Analysis (PCA) showed the global *Mycobacterial tuberculosis* clinical isolates and laboratory H37Rv strain showed differences in their metabolite composition.

**Supplementary Table 1:** Details of the reported Mycobacterium tuberculosis strains and the newly identified clinical isolates used for comparison.

| **Organism Names** | **Type** | **lineage** | **Drug Susceptible/ Resistance (S/R)** | **Sequenced by** | **Isolated place** | **NCBI ID** | **Tax Id** | **Length (in bp)** | **Pubmed ID of references** |
| --- | --- | --- | --- | --- | --- | --- | --- | --- | --- |
| *M. tuberculosis* **H37Rv** | Reference | Euro American |  | Sangers Institute |  | [AL123456](https://www.ncbi.nlm.nih.gov/nuccore/AL123456) | [83332](https://www.ncbi.nlm.nih.gov/Taxonomy/Browser/wwwtax.cgi?mode=Info&id=83332) | 44,11,532 | [9634230](https://www.ncbi.nlm.nih.gov/pubmed/9634230) |
| *M. tuberculosis* **H37Ra** |  |  |  | CNHG, Shanghai |  | [CP000611](https://www.ncbi.nlm.nih.gov/nuccore/CP000611) | [419947](https://www.ncbi.nlm.nih.gov/Taxonomy/Browser/wwwtax.cgi?mode=Info&id=419947) | 44,19,977 | [18584054](https://www.ncbi.nlm.nih.gov/pubmed/18584054) |
| *M. tuberculosis* **CDC1551** |  |  |  | TIGR | Tennessee and Kentucky, US | [AE000516](https://www.ncbi.nlm.nih.gov/nuccore/AE000516) | [83331](https://www.ncbi.nlm.nih.gov/Taxonomy/Browser/wwwtax.cgi?mode=Info&id=83331) | 44,03,837 | [12218036](https://www.ncbi.nlm.nih.gov/pubmed/12218036) |
| *M. tuberculosis* **ATCC 35801 / Erdman** | Laboratory strain |  |  | NCGM, Japan | Mayo Clinic, Minnesota | [AP012340](https://www.ncbi.nlm.nih.gov/nuccore/AP012340) | [652616](https://www.ncbi.nlm.nih.gov/Taxonomy/Browser/wwwtax.cgi?mode=Info&id=652616) | 43,92,353 | [22535945](https://www.ncbi.nlm.nih.gov/pubmed/22535945) |
| *M. tuberculosis* **S1** | Clinical strain | East African Indian (EAI) |  | ICGEB, Delhi, India and it’s collaborator | Agartala, Tripura, India |  |  | 43,99,078 |  |
| *M. tuberculosis* **S4** |  |  | R |  |  |  |  | 43,96,813 |  |
| *M. tuberculosis* **S5** |  |  | R |  |  |  |  | 44,56,211 |  |
| *M. tuberculosis* **S6** |  |  | R |  |  |  |  | 44,01,104 |  |
| *M. tuberculosis* **S7** |  |  | R |  |  | [ASM982363](https://www.ncbi.nlm.nih.gov/assembly/GCF_009823635.1/) | [1773](https://www.ncbi.nlm.nih.gov/Taxonomy/Browser/wwwtax.cgi?id=1773) | 43,91,229 |  |
| *M. tuberculosis* **S10** |  |  | R |  |  |  |  | 44,05,137 |  |
| *M. tuberculosis* **RGTB 327** |  |  |  | RGTB, Hyderabad | Kerala, India | [CP003233](https://www.ncbi.nlm.nih.gov/nuccore/CP003233) | [1091500](https://www.ncbi.nlm.nih.gov/Taxonomy/Browser/wwwtax.cgi?mode=Info&id=1091500) | 43,80,119 | [22843573](https://www.ncbi.nlm.nih.gov/pubmed/22843573) |
| *M. tuberculosis* **RGTB 423** |  |  |  |  |  | [CP003234](https://www.ncbi.nlm.nih.gov/nuccore/CP003234) | [1091501](https://www.ncbi.nlm.nih.gov/Taxonomy/Browser/wwwtax.cgi?mode=Info&id=1091501) | 44,06,587 | [22843573](https://www.ncbi.nlm.nih.gov/pubmed/22843573) |
| *M. tuberculosis* **NITR 206 / EAI5** |  |  |  | NITR, Chennai | Tamilnadu, India | [CP005387](https://www.ncbi.nlm.nih.gov/nuccore/CP005387) | [1310115](https://www.ncbi.nlm.nih.gov/Taxonomy/Browser/wwwtax.cgi?mode=Info&id=1310115) | 43,90,306 | [23788533](https://www.ncbi.nlm.nih.gov/pmc/articles/PMC3707582/) |
| *M. tuberculosis* **EAI5** |  |  |  | Mumbai University, Mumbai | Mumbai, India | [CP006578](https://www.ncbi.nlm.nih.gov/nuccore/CP006578) | [1306414](https://www.ncbi.nlm.nih.gov/Taxonomy/Browser/wwwtax.cgi?mode=Info&id=1306414) | 43,91,174 | [24604653](https://www.ncbi.nlm.nih.gov/pubmed/24604653) |
| *M. tuberculosis* **F11** |  | Euro American |  | Broad Institute | Western Cape of South Africa | [CP000717](https://www.ncbi.nlm.nih.gov/nuccore/CP000717) | [336982](https://www.ncbi.nlm.nih.gov/Taxonomy/Browser/wwwtax.cgi?mode=Info&id=336982) | 44,24,435 |  |
| *M. tuberculosis* **KZN 1435** |  |  |  |  | KwaZulu-Natal, South Africa. | [CP001658](https://www.ncbi.nlm.nih.gov/nuccore/CP001658) | [478434](https://www.ncbi.nlm.nih.gov/Taxonomy/Browser/wwwtax.cgi?mode=Info&id=478434) | 43,98,250 |  |
| *M. tuberculosis* **KZN 4207** |  |  |  |  |  | [CP001662](https://www.ncbi.nlm.nih.gov/nuccore/CP001662) | [478433](https://www.ncbi.nlm.nih.gov/Taxonomy/Browser/wwwtax.cgi?mode=Info&id=478433) | 43,94,985 |  |
| *M. tuberculosis* **KZN 605** |  |  |  |  |  | [CP001976](https://www.ncbi.nlm.nih.gov/nuccore/CP001976) | [478435](https://www.ncbi.nlm.nih.gov/Taxonomy/Browser/wwwtax.cgi?mode=Info&id=478435) | 43,99,120 |  |
| *M. tuberculosis* **CTRI-2** |  |  |  | SRI PCM | Central region European Russia | [CP002992](https://www.ncbi.nlm.nih.gov/nuccore/CP002992) | [707235](https://www.ncbi.nlm.nih.gov/Taxonomy/Browser/wwwtax.cgi?mode=Info&id=707235) | 43,98,525 | [23437175](https://www.ncbi.nlm.nih.gov/pubmed/23437175) |
| *M. tuberculosis* **UT 205** |  |  |  | CNSG, CCITB | Colombian clinical isolate | [HE608151](https://www.ncbi.nlm.nih.gov/nuccore/HE608151) | [1097669](https://www.ncbi.nlm.nih.gov/Taxonomy/Browser/wwwtax.cgi?mode=Info&id=1097669) | 44,18,088 | [22404577](https://www.ncbi.nlm.nih.gov/pubmed/22404577) |
| *M. tuberculosis* **NITR 202 / Haarlem** |  |  |  | NITR, Chennai | Tamilnadu, India | [CP004886](https://www.ncbi.nlm.nih.gov/nuccore/CP004886) | [1304279](https://www.ncbi.nlm.nih.gov/Taxonomy/Browser/wwwtax.cgi?mode=Info&id=1304279) | 44,04,786 | [23788533](https://www.ncbi.nlm.nih.gov/pmc/articles/PMC3707582/) |
| *M. tuberculosis* **7199-99** |  |  |  | Bielefeld University |  | [HE663067](https://www.ncbi.nlm.nih.gov/nuccore/HE663067) | [1138877](https://www.ncbi.nlm.nih.gov/Taxonomy/Browser/wwwtax.cgi?mode=Info&id=1138877) | 44,21,197 | [23424287](https://www.ncbi.nlm.nih.gov/pubmed/23424287) |
| *M. tuberculosis* **Haarlem** |  |  | S | Broad Institute | Netherland | [CP001664](https://www.ncbi.nlm.nih.gov/nuccore/CP001664) | [395095](https://www.ncbi.nlm.nih.gov/Taxonomy/Browser/wwwtax.cgi?mode=Info&id=395095) | 44,08,224 |  |
| *M. tuberculosis* **NITR 204 / CAS** |  | Central Asian |  | NITR, Chennai | Tamilnadu, India | [CP005386](https://www.ncbi.nlm.nih.gov/nuccore/CP005386) | [1310114](https://www.ncbi.nlm.nih.gov/Taxonomy/Browser/wwwtax.cgi?mode=Info&id=1310114) | 43,92,876 | 23788533 |
| *M. tuberculosis* **NITR 203 / Beijing** |  | East Asian/  Beijing | S |  |  | [CP005082](https://www.ncbi.nlm.nih.gov/nuccore/CP005082) | [1306400](https://www.ncbi.nlm.nih.gov/Taxonomy/Browser/wwwtax.cgi?mode=Info&id=1306400) | 44,11,128 | 23788533 |
| *M. tuberculosis* **CCDC 5079** |  |  | R | CCDCP | Fujian Province, China | [CP001641](https://www.ncbi.nlm.nih.gov/nuccore/CP001641) | [443149](https://www.ncbi.nlm.nih.gov/Taxonomy/Browser/wwwtax.cgi?mode=Info&id=443149) | 43,98,812 | [21914894](https://www.ncbi.nlm.nih.gov/pubmed/21914894) |
| *M. tuberculosis* **CCDC 5180** |  |  | R |  |  | [CP001642](https://www.ncbi.nlm.nih.gov/nuccore/CP001642) | [443150](https://www.ncbi.nlm.nih.gov/Taxonomy/Browser/wwwtax.cgi?mode=Info&id=443150) | 44,05,981 | [21914894](https://www.ncbi.nlm.nih.gov/pubmed/21914894) |
| *M. tuberculosis* **BT1** |  |  | S | CUHK, Hong Kong | Hong Kong | [CP002883](https://www.ncbi.nlm.nih.gov/nuccore/CP002883) | [1010836](https://www.ncbi.nlm.nih.gov/Taxonomy/Browser/wwwtax.cgi?mode=Info&id=1010836) | 43,99,405 |  |
| *M. tuberculosis* **BT2** |  |  | S |  |  | [CP002882](https://www.ncbi.nlm.nih.gov/nuccore/CP002882) | [1010835](https://www.ncbi.nlm.nih.gov/Taxonomy/Browser/wwwtax.cgi?mode=Info&id=1010835) | 44,01,899 |  |
| *M. tuberculosis* **HKBS1** |  |  | R |  |  | [CP002871](https://www.ncbi.nlm.nih.gov/nuccore/CP002871) | [1010834](https://www.ncbi.nlm.nih.gov/Taxonomy/Browser/wwwtax.cgi?mode=Info&id=1010834) | 44,07,929 |  |

**Supplementary Table 2:** Genome features of new clinical Mycobacterium tuberculosis isolates. ANI: Average Nucleotide Identity, SNPs: Single nucleotide polymorphisms.

|  | **H37Rv** | **S1** | **S4** | **S5** | **S6** | **S7** | **S10** |
| --- | --- | --- | --- | --- | --- | --- | --- |
| **Size (in bp)** | 44,19,977 | 43,99,078 | 43,96,813 | 44,56,211 | 44,01,104 | 43,91,229 | 44,05,137 |
| **Taxonomy ID** | 419947 | NA | NA | NA | NA | 1773 | NA |
| **Coverage** | NA | 99x | 99x | 99x | 99x | 99x | 99x |
| **GC content** | 65.6 | 65.6 | 65.6 | 65.6 | 65.6 | 65.6 | 65.5 |
| **N50** | NA | 115355 | 101973 | 167008 | 119047 | 129971 | 106868 |
| **L50** | NA | 12 | 12 | 10 | 12 | 11 | 13 |
| **Contigs** | 1 | 111 | 202 | 306 | 210 | 145 | 124 |
| **ANI** | 100% | 99.72 | 99.73 | 99.72 | 99.77 | 99.79 | 99.75 |
| **SNPs** | NA | 2494 | 2519 | 3148 | 1950 | 1280 | 2870 |

**Supplementary Table 3:** SNPs annotated from genes associated with drug resistance among the new clinical Mycobacterium tuberculosis isolates. The presence single nucleotide polymorphisms (SNPs) is represented by green and absence in red colour.

| **Genes** | **Drugs** | **SNPs** | **S1** | **S4** | **S5** | **S6** | **S7** | **S10** |
| --- | --- | --- | --- | --- | --- | --- | --- | --- |
| gyrA | Fluroquinolone | E21Q |  |  |  |  |  |  |
|  |  | S95T |  |  |  |  |  |  |
|  |  | A384V |  |  |  |  |  |  |
|  |  | G668D |  |  |  |  |  |  |
|  |  | L751V |  |  |  |  |  |  |
| gyrB |  | G92S |  |  |  |  |  |  |
|  |  | M291I |  |  |  |  |  |  |
| iniA | Isoniazid | H481Q |  |  |  |  |  |  |
| katG promoter |  | S315T |  |  |  |  |  |  |
|  |  | R463L |  |  |  |  |  |  |
|  |  | A234G |  |  |  |  |  |  |
|  |  | A431V |  |  |  |  |  |  |
|  |  | G300W |  |  |  |  |  |  |
| fabG1-promoter |  | -15 C>T in nucleotide |  |  |  |  |  |  |
| ahpC-promoter |  | -142 G>A in nucleotide |  |  |  |  |  |  |
| rpoC | Rifampicin | A172V |  |  |  |  |  |  |
|  |  | L516P |  |  |  |  |  |  |
|  |  | A621T |  |  |  |  |  |  |
|  |  | P601L |  |  |  |  |  |  |
| rpoB |  | D516G |  |  |  |  |  |  |
|  |  | S450L |  |  |  |  |  |  |
|  |  | H526T |  |  |  |  |  |  |
|  |  | L511R |  |  |  |  |  |  |
| embC | Ethambutol | T270I |  |  |  |  |  |  |
|  |  | N394D |  |  |  |  |  |  |
| embB |  | Q497R |  |  |  |  |  |  |
|  |  | Q853P |  |  |  |  |  |  |
|  |  | E378A |  |  |  |  |  |  |
|  |  | T1082 |  |  |  |  |  |  |
| embA- promoter |  | V206M |  |  |  |  |  |  |
|  |  | P913S |  |  |  |  |  |  |
| embC-promoter |  | -141 G>A in nucleotide |  |  |  |  |  |  |
| embR |  | C110Y |  |  |  |  |  |  |
| ethA-promoter | Ethionamide | N345K |  |  |  |  |  |  |
| ethR |  | A9T |  |  |  |  |  |  |
| gidB | Aminoglycoside | W123C |  |  |  |  |  |  |
| tlyA |  | Insertion 180insP |  |  |  |  |  |  |
| 16s rRNA (rrs) |  | A503C |  |  |  |  |  |  |
|  |  | A514C |  |  |  |  |  |  |
| 23s rRNA |  | A2273G |  |  |  |  |  |  |
| rpsL |  | K88R |  |  |  |  |  |  |
| drrA |  | H309D |  |  |  |  |  |  |
| ubiA |  | E149D |  |  |  |  |  |  |
| murA |  | C117D |  |  |  |  |  |  |

**Supplementary Table 4:** List of genes associated with anti-TB drug as their targets or drug modifying proteins that provide resistance to MDR and XDR strains.

| **Drug class** | **Drug associated enzymes/transporters** | **Mechanism of resistance** |
| --- | --- | --- |
| Rifamycin | rpoB | Drug target alteration |
|  | rpoC |  |
|  | RbpA | Antibiotic target protection |
| Isoniazid | iniA | Mutation at drug target site |
|  | AhpC | Drug alteration or reduced activation |
|  | KatG |  |
|  | fabG1 | Mutation at drug target site |
| Ethambutol | embA | Mutation prevents drug binding |
|  | embB |  |
|  | embC |  |
|  | embR | Drug alteration |
|  | iniA | Mutation at drug target site |
| Aminoglycoside | RpsL | Mutation at drug target site |
|  | 16s rRNA of ribosome |  |
|  | 23s rRNA of ribosome |  |
|  | TlyA |  |
|  | AAC (2')-lc | Inactivation of drug by modification |
|  | gidB | Methylation of drug target site |
| Fluoroquinolone | GyrA | Drug target mutation |
|  | GyrB |  |
|  | mfpA | Antibiotic efflux |
|  | Rv2686c | Fluoroquinolone drug transporters |
|  | Rv2687c |  |
|  | Rv2688c |  |
| Lincomycin | Erm (37) | Methylation of drug target site |
| Macrolide |  |  |
| Macrolide | 23s rRNA of ribosome | Mutation at drug target sites |
| Thioamides | EthR | Repression of EthA activity |
|  | EthA | Mutation leads to lack of drug activation |
|  | fabG1 |  |

**Supplementary Table 5:** Categorization of annotated genes from the reference Mycobacterial tuberculosis strains (H37Ra and H37Rv) and new clinical isolates.

| **Complete genome annotation Statistics** | | | | | | | | |
| --- | --- | --- | --- | --- | --- | --- | --- | --- |
|  | **H37Ra** | **H37Rv** | **S1** | **S4** | **S5** | **S6** | **S7** | **S10** |
| **CDS** | 4309 | 4299 | 4347 | 4425 | 4551 | 4439 | 4336 | 4385 |
| **tRNAs** | 45 | 45 | 45 | 45 | 45 | 45 | 45 | 45 |
| **rRNAs** | 2 | 2 | 2 | 2 | 2 | 2 | 2 | 2 |
| **Genes involved in Subsystem features** | 995 | 989 | 1007 | 1023 | 1048 | 1014 | 1002 | 1001 |
| **Subsystem features** | **Feature counts** | | | | | | | |
| Cofactors, Vitamins, Prosthetic Groups, Pigments | 142 | 142 | 145 | 147 | 153 | 141 | 149 | 143 |
| Cell Wall and Capsule | 27 | 27 | 27 | 28 | 28 | 29 | 27 | 27 |
| Virulence, Disease and Defence | 45 | 43 | 53 | 55 | 55 | 50 | 42 | 46 |
| Potassium metabolism | 7 | 7 | 7 | 7 | 7 | 7 | 7 | 7 |
| Miscellaneous | 23 | 23 | 23 | 23 | 24 | 23 | 23 | 23 |
| Phage, Prophages, Transposable elements, Plasmids | 2 | 3 | 2 | 1 | 2 | 2 | 3 | 2 |
| Membrane Transport | 25 | 25 | 25 | 25 | 25 | 26 | 25 | 25 |
| Iron acquisition and metabolism | 2 | 2 | 2 | 2 | 3 | 2 | 2 | 2 |
| RNA Metabolism | 45 | 34 | 45 | 45 | 47 | 45 | 45 | 45 |
| Nucleosides and Nucleotides | 70 | 70 | 68 | 68 | 74 | 68 | 70 | 68 |
| Protein Metabolism | 165 | 165 | 166 | 168 | 169 | 168 | 165 | 166 |
| Regulation and Cell signalling | 99 | 99 | 100 | 101 | 101 | 102 | 100 | 100 |
| Secondary Metabolism | 1 | 1 | 1 | 1 | 1 | 1 | 1 | 1 |
| DNA Metabolism | 92 | 92 | 91 | 93 | 96 | 86 | 95 | 91 |
| Fatty Acids, Lipids, and Isoprenoids | 141 | 141 | 141 | 144 | 145 | 149 | 145 | 140 |
| Nitrogen Metabolism | 20 | 20 | 20 | 21 | 20 | 20 | 20 | 20 |
| Dormancy and Sporulation | 1 | 1 | 1 | 1 | 1 | 1 | 1 | 1 |
| Respiration | 73 | 73 | 73 | 75 | 75 | 73 | 73 | 73 |
| Stress Response | 26 | 26 | 26 | 27 | 27 | 26 | 26 | 26 |
| Metabolism of Aromatic Compounds | 7 | 7 | 9 | 8 | 7 | 7 | 7 | 7 |
| Amino Acids and Derivatives | 265 | 265 | 274 | 278 | 306 | 265 | 267 | 275 |
| Sulfur Metabolism | 7 | 7 | 7 | 7 | 7 | 7 | 8 | 7 |
| Phosphorus Metabolism | 27 | 27 | 27 | 27 | 29 | 27 | 27 | 27 |
| Carbohydrates | 199 | 198 | 195 | 198 | 203 | 210 | 198 | 196 |

**Supplementary Table 6:** List of additionally annotated proteins in multidrug resistance strains of Mtb that might having functions related to drug resistance and other metabolites utilization.

| **Clinical**  **isolate** | **Category** | **Subcategory** | **Function** | **Quantity** |
| --- | --- | --- | --- | --- |
| **S1** | Cofactors, Vitamins, Prosthetic Groups, Pigments | Biotin biosynthesis | Long-chain-fatty-acid--CoA ligase (EC 6.2.1.3) | 2 |
|  |  | Folate and pterions | Fumarylacetoacetate hydrolase family protein | 1 |
|  | Virulence, Disease and Defence | Invasion and intracellular resistance | ESAT-6-like protein EsxK | 3 |
|  |  |  | ESAT-6-like protein EsxL | 7 |
|  | Protein Metabolism | Protein degradation | Prolyl endopeptidase (EC 3.4.21.26) | 1 |
|  | Regulation and Cell signalling | Programmed Cell Death and Toxin-antitoxin Systems | Cell division protein DivIC (FtsB), stabilizes FtsL against RasP cleavage | 1 |
|  | Amino Acids and Derivatives | Leucine Biosynthesis | 2-isopropylmalate synthase (EC 2.3.3.13) | 1 |
|  |  | Branched-Chain Amino Acid Biosynthesis | 3-isopropylmalate dehydrogenase (EC 1.1.1.85) | 1 |
| **S4** | Amino Acids and Derivatives | Branched-Chain Amino Acid Biosynthesis | 3-isopropylmalate dehydrogenase (EC 1.1.1.85) | 1 |
|  | DNA Metabolism | DNA Repair Base Excision | DNA ligase (EC 6.5.1.2) | 1 |
|  |  | DNA uptake, competence | FIG000557: hypothetical protein co-occurring with RecR | 1 |
|  | RNA Metabolism | tRNA modification Bacteria | tRNA uridine 5-carboxymethylaminomethyl modification enzyme GidA | 1 |
|  | Stress Response | Glutathione: Non-redox reactions | Glutathione S-transferase (EC 2.5.1.18) | 1 |
|  | Virulence, Disease and Defence | Copper homeostasis | Copper resistance protein B | 1 |
|  |  | Invasion and intracellular resistance | ESAT-6-like protein EsxK | 5 |
|  |  | Invasion and intracellular resistance | ESAT-6-like protein EsxL | 6 |
|  | Cell Wall and Capsule | Murein Hydrolases | D-alanyl-D-alanine carboxypeptidase (EC 3.4.16.4) | 1 |
|  | Cofactors, Vitamins, Prosthetic Groups, Pigments | Biotin biosynthesis | Biotin synthase (EC 2.8.1.6) | 1 |
|  |  |  | Long-chain-fatty-acid--CoA ligase (EC 6.2.1.3) | 3 |
|  | Protein Metabolism | Protein degradation | D-alanyl-D-alanine carboxypeptidase (EC 3.4.16.4) | 1 |
|  |  |  | Prolyl endopeptidase (EC 3.4.21.26) | 1 |
|  | Regulation and Cell signalling | Possible new toxin-antitoxin system including DivIC | Cell division protein DivIC (FtsB), stabilizes FtsL against RasP cleavage | 1 |
|  |  | Toxin-Antitoxin MT1 | Toxin 1, PIN domain | 1 |
|  | Fatty Acids, Lipids, and Isoprenoids | Acyl-CoA thioesterase II | TesB-like acyl-CoA thioesterase 3 | 1 |
|  |  | Glycerolipid and Glycerophospholipid Metabolism in Bacteria | Aldehyde dehydrogenase (EC 1.2.1.3) | 1 |
|  | Stress Response | Glutathione: Non-redox reactions | Glutathione S-transferase (EC 2.5.1.18) | 1 |
|  | Amino Acids and Derivatives | Aromatic amino acids and derivatives | Chorismate synthase (EC 4.2.3.5) | 1 |
|  |  | Leucine Biosynthesis | 2-isopropylmalate synthase (EC 2.3.3.13) | 1 |
|  |  | Leucine Degradation and HMG-CoA Metabolism | Succinyl-CoA:3-ketoacid-coenzyme A transferase subunit A (EC 2.8.3.5) | 1 |
|  |  | Glutamine, Glutamate, Aspartate and Asparagine Biosynthesis | Glutamate synthase [NADPH] large chain (EC 1.4.1.13) | 1 |
|  | Respiration | Electron accepting reactions | Ferredoxin reductase | 1 |
|  |  | Quinone oxidoreductase family | Quinone oxidoreductase (EC 1.6.5.5) | 1 |
| **S5** | Amino Acids and Derivatives | Arginine; urea cycle, polyamines | Acetylglutamate kinase (EC 2.7.2.8) | 1 |
|  |  | Branched-Chain Amino Acid Biosynthesis | 3-isopropylmalate dehydrogenase (EC 1.1.1.85) | 1 |
|  |  | Isoleucine degradation | Probable acyl-CoA dehydrogenase (EC 1.3.99.3) | 1 |
|  | Carbohydrates | Chitin and N-acetylglucosamine utilization | Chitinase (EC 3.2.1.14) | 1 |
|  |  | Maltose and Maltodextrin Utilization | Glucoamylase (EC 3.2.1.3) | 1 |
|  | Clustering-based subsystems | ADP-phosphoribose and NAD-dependent acetylation | Aspartate aminotransferase (EC 2.6.1.1) | 1 |
|  |  | tRNA-methylthiotransferase containing cluster | Apolipoprotein N-acyltransferase (EC 2.3.1.-) | 1 |
|  | DNA Metabolism | DNA Repair Base Excision | DNA ligase (EC 6.5.1.2) | 1 |
|  |  | DNA repair, bacterial RecBCD pathway | Protease III precursor (EC 3.4.24.55) | 1 |
|  |  | Restriction-Modification System | Type III restriction-modification system methylation subunit (EC 2.1.1.72) | 1 |
|  | Fatty Acids, Lipids, and Isoprenoids | Fatty Acid Biosynthesis FASII | Acyl carrier protein phosphodiesterase (EC 3.1.4.14) | 1 |
|  | RNA Metabolism | tRNA modification Bacteria | tRNA uridine 5-carboxymethylaminomethyl modification enzyme GidA | 1 |
|  | Stress Response | Bacterial hemoglobins | diguanylate cyclase/phosphodiesterase (GGDEF & EAL domains) with PAS/PAC sensor(s) | 1 |
|  | Virulence, Disease and Defense | Copper homeostasis: copper tolerance | Copper homeostasis protein CutE | 1 |
|  | Cofactors, Vitamins, Prosthetic Groups, Pigments | Biotin biosynthesis | Long-chain-fatty-acid--CoA ligase (EC 6.2.1.3) | 3 |
|  |  | Folate and pterines | Dihydrolipoamide acetyltransferase component of pyruvate dehydrogenase complex (EC 2.3.1.12) | 1 |
|  |  |  | Phosphoribosylaminoimidazolecarboxamide formyl transferase (EC 2.1.2.3) | 1 |
|  |  |  | Serine hydroxymethyl transferase (EC 2.1.2.1) | 1 |
|  |  | Folate Biosynthesis | Thymidylate synthase (EC 2.1.1.45) | 1 |
|  |  |  | FIG027937: secreted protein | 1 |
|  |  | Folate biosynthesis cluster | transmembrane protein, distant homology with ydbS | 1 |
|  |  | Pyridoxin (Vitamin B6) Biosynthesis | D-3-phosphoglycerate dehydrogenase (EC 1.1.1.95) | 1 |
|  | Cell Wall and Capsule | Rhamnose containing glycans | UDP-glucose 4-epimerase (EC 5.1.3.2) | 1 |
|  | Virulence, Disease and Defence | Invasion and intracellular resistance | Translation elongation factor Tu | 1 |
|  |  |  | ESAT-6-like protein EsxK | 4 |
|  |  |  | ESAT-6-like protein EsxL | 4 |
|  | Amino Acids and Derivatives | Alanine biosynthesis | Cysteine desulfurase (EC 2.8.1.7) | 1 |
|  |  | Glycine Biosynthesis | Serine hydroxymethyl transferase (EC 2.1.2.1) | 1 |
|  |  | Glycine and Serine Utilization | D-3-phosphoglycerate dehydrogenase (EC 1.1.1.95) | 1 |
|  |  |  | Serine hydroxymethyl transferase (EC 2.1.2.1) | 1 |
|  |  | Leucine Biosynthesis | 2-isopropylmalate synthase (EC 2.3.3.13) | 1 |
|  |  | Valine degradation | Methylmalonate-semialdehyde dehydrogenase (EC 1.2.1.27) | 1 |
|  |  | Methionine Biosynthesis | 5-methyltetrahydrofolate--homocysteine methyltransferase (EC 2.1.1.13) | 1 |
|  | Phosphorus Metabolism | Phosphate metabolism | Exopolyphosphatase (EC 3.6.1.11) | 1 |
| **S6** | Carbohydrates | Fermentations: Mixed acid | Phosphoenolpyruvate carboxylase (EC 4.1.1.31) | 1 |
|  | Clustering-based subsystems | Stress related cluster | Arsenical pump-driving ATPase (EC 3.6.3.16) | 1 |
|  | DNA Metabolism | DNA Repair Base Excision | DNA ligase (EC 6.5.1.2) | 1 |
|  |  | DNA processing cluster | FIG000557: hypothetical protein co-occurring with RecR | 1 |
|  | RNA Metabolism | tRNA modification Bacteria | tRNA uridine 5-carboxymethylaminomethyl modification enzyme GidA | 1 |
|  | Cell Wall and Capsule | Lipid-linked oligosaccharide synthesis related cluster | Apolipoprotein N-acyltransferase (EC 2.3.1.-) in lipid-linked oligosaccharide synthesis cluster | 1 |
|  |  | Murein Hydrolases | D-alanyl-D-alanine carboxypeptidase (EC 3.4.16.4) | 1 |
|  | Virulence, Disease and Defence | Invasion and intracellular resistance | ESAT-6-like protein EsxK | 3 |
|  |  |  | ESAT-6-like protein EsxL | 3 |
|  |  | Copper homeostasis | Copper-translocating P-type ATPase (EC 3.6.3.4) | 1 |
|  | Membrane Transport | Copper Transport System | Copper-translocating P-type ATPase (EC 3.6.3.4) | 1 |
|  | Regulation and Cell signalling | Toxin-Antitoxin MT1 | Toxin 1, PIN domain | 1 |
|  |  | cAMP signalling in bacteria | Adenylate cyclase (EC 4.6.1.1) | 2 |
|  | Fatty Acids, Lipids, and Isoprenoids | Fatty Acid Biosynthesis FASII | Biotin carboxyl carrier protein of acetyl-CoA carboxylase | 1 |
|  |  |  | Biotin carboxylase of acetyl-CoA carboxylase (EC 6.3.4.14) | 1 |
|  |  | Carotenoids, Isoprenoids for Quinones and Polyprenyl Diphosphate Biosynthesis | (2E,6E)-farnesyl diphosphate synthase (EC 2.5.1.10) | 1 |
|  |  | Nonmevalonate Branch of Isoprenoid Biosynthesis | 2-C-methyl-D-erythritol 4-phosphate cytidylyltransferase (EC 2.7.7.60) | 1 |
|  |  | Glycerolipid and Glycerophospholipid Metabolism in Bacteria | Glycerol-3-phosphate acyltransferase (EC 2.3.1.15) | 2 |
|  | Carbohydrates | Central carbohydrate metabolism | Dihydrolipoamide acyltransferase component of branched-chain alpha-keto acid dehydrogenase complex (EC 2.3.1.168) | 1 |
|  |  | Pyruvate Alanine Serine Interconversions | Omega-amino acid--pyruvate aminotransferase (EC 2.6.1.18) | 2 |
|  |  | Mannose Metabolism | Alpha-1,2-mannosidase | 1 |
| **S7** | Amino Acids and Derivatives | Arginine; urea cycle, polyamines | Putrescine ABC transporter putrescine-binding protein PotF (TC 3.A.1.11.2) | 1 |
|  | RNA Metabolism | tRNA modification Bacteria | tRNA uridine 5-carboxymethylaminomethyl modification enzyme GidA | 1 |
|  | Cofactors, Vitamins, Prosthetic Groups, Pigments | Biotin biosynthesis | Long-chain-fatty-acid--CoA ligase (EC 6.2.1.3) | 2 |
|  |  | Folate and pterines | Pterin-4-alpha-carbinolamine dehydratase (EC 4.2.1.96) | 1 |
|  |  | Pyridoxin (Vitamin B6) Biosynthesis and Thiamin biosynthesis | 1-deoxy-D-xylulose 5-phosphate synthase (EC 2.2.1.7) | 2 |
|  | DNA Metabolism | DNA Repair Base Excision | ATP-dependent DNA ligase (EC 6.5.1.1) | 1 |
|  |  | DNA repair, bacterial | A/G-specific adenine glycosylase (EC 3.2.2.-) | 1 |
|  |  | Leucine Biosynthesis | 3-isopropylmalate dehydratase large subunit (EC 4.2.1.33) | 1 |
| **S10** | Amino Acids and Derivatives | Branched-Chain Amino Acid Biosynthesis | 3-isopropylmalate dehydrogenase (EC 1.1.1.85) |  |
|  | RNA Metabolism | tRNA modification Bacteria | tRNA uridine 5-carboxymethylaminomethyl modification enzyme GidA |  |
|  | Virulence, Disease and Defence | Invasion and intracellular resistance | ESAT-6-like protein EsxL | 3 |
|  | Amino Acids and Derivatives | Putrescine utilization pathways | Gamma-aminobutyrate:alpha-ketoglutarate aminotransferase (EC 2.6.1.19) |  |
